# The autumn COVID-19 surge dates in Europe are linked to latitudes and not to temperature, nor to humidity, pointing vitamin D as a contributing factor

**DOI:** 10.1101/2020.10.28.20221176

**Authors:** Stephan Walrand

## Abstract

**Purpose:** Determining the triggering factor of the sudden surge of the daily new COVID-19 cases arising in most European countries during 2020 Autumn.

**Methods:** The dates of the surge were determined using a fitting of the two last months reported daily new cases in 18 European countries of latitude ranging from 39° to 62°.

**Results:** The study proves no correlation between the country surge date and its 2 weeks preceding temperature or humidity, but shows an impressive linear correlation with its latitude. The country surge date corresponds to the time when its sun UV daily dose drops below ≈ 34% of that of 0° latitude. Introducing reported seasonal blood 25-hydroxyvitamin D (25(OH)D) concentration variation into reported link between acute respiratory track infection risk with 25(OH)D concentration quantitatively explains the surge dynamics.

**Conclusions:** Several studies already substantiated a 25(OH)D concentration impact on COVID-19 severity. However by comparing different patients populations, discriminate whether low 25(OH)D concentration is a real factor of covid-19 severity or only a marker of another weakness being the primary severity factor can be challenging. The date of the surge is an intrapopulation observation and has the benefit to be only triggered by a parameter globally affecting the population, i.e. the sun UV daily dose decreases. The results support that low 25(OH)D concentration is thus well a contributing factor of COVID-19 severity, which joined with the previous studies makes a convincing bundle of evidence

## Introduction

Most European countries underwent in autumn an unexpected surge of the daily new COVID-19 cases (Fig. 1), imposing in emergency new confinement rules and lockdowns.

**Figure 1:**
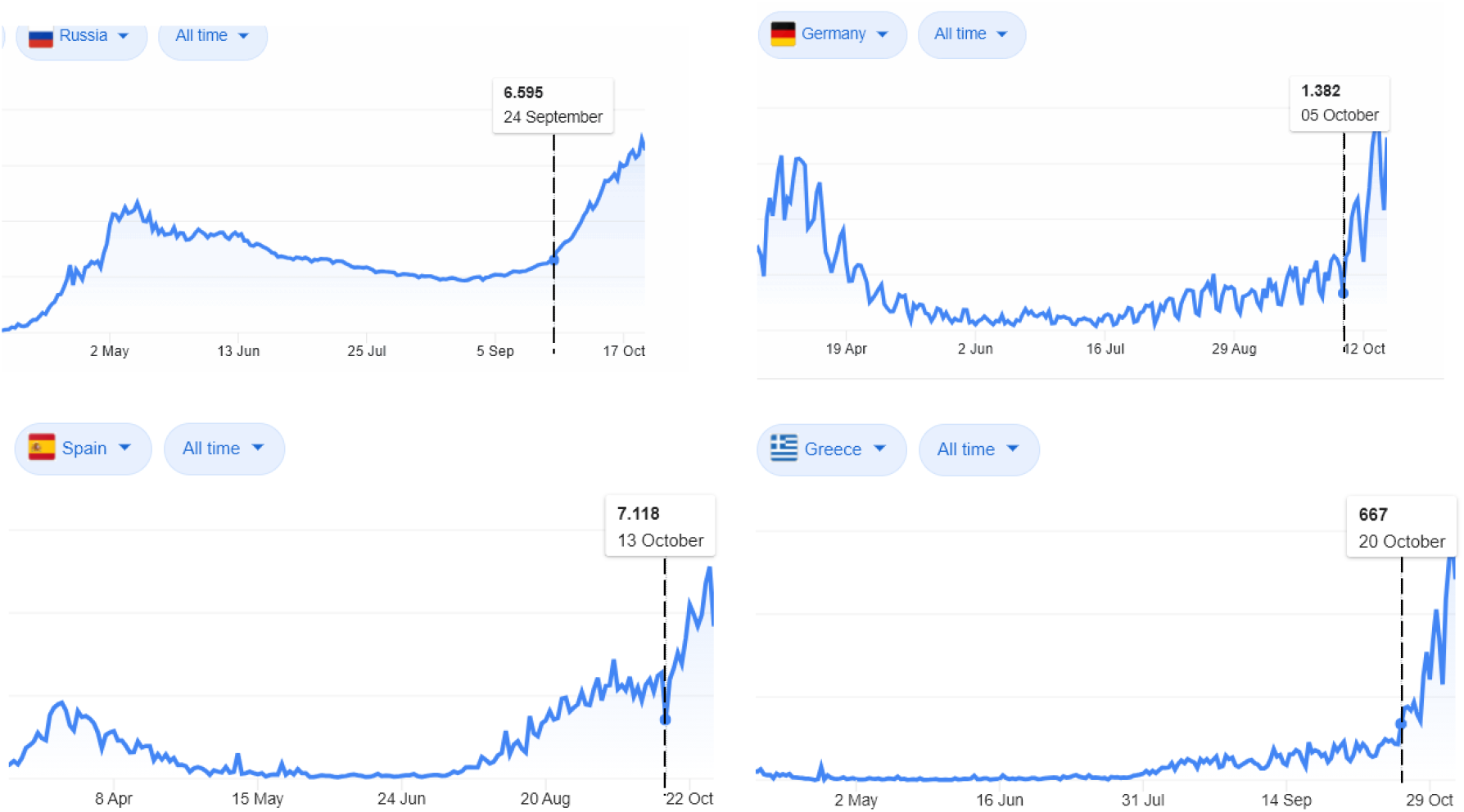
Typical examples of daily new COVID-19 cases (extracted from the Google home page when searching “covid”). All curves exhibit a clear surge of the growing rates.

A common reported explanation is the decreasing temperature. The aim of this study is to challenge this assumption against a pure latitude impact.

## Material and method

### Data sources

The countries daily new COVID-19 cases were obtained from the European Union agency European Centre for Disease Prevention and Control (https://www.ecdc.europa.eu/en/publications-data/download-todays-data-geographic-distribution-covid-19-cases-worldwide).

The country population weighted center (PWC) latitudes were obtained from the Baylor University population resource (http://cs.ecs.baylor.edu/~hamerly/software/europe_population_weighted_centers.txt)

The averaged 2 weeks temperatures and humidity preceding the surge dates were computed from https://rp5.ru which collects the archives of all the airport weather stations in the world. For each country, an airport close to the PWC was chosen (see supplementary excel file). Averaged temperature and humidity were computed between 8h00 to 20h00, outside this period the population being mostly indoor.

School opening dates of 15 countries on the 18 ones studied were found at: https://eacea.ec.europa.eu/national-policies/eurydice/sites/eurydice/files/school_calendar_2020_21_0.pdf

The theoretical sun UVB daily dose for vitamin D production, as a function of latitude and of the day of the year, was derived from the digitalization of fig. 1B in [1].

All these data are in the supplementary excel file.

### Surge date determination

The date of the surge was automatically determined by fitting the two last months of the daily new COVID-19 cases with the empirical model:

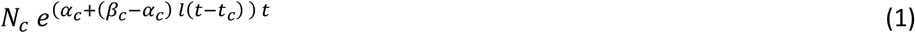

where *l* is the logistic function:

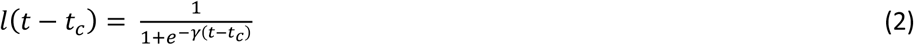

*t*_*c*_ is the date when the exponential coefficient, coming from the initial value *α*_*c*_, crosses the value 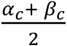 before tending towards *β*_c_ when *t* → ∞. *γ* is the steepness of this changing. The date of the surge was defined as the time when 10% of 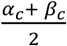 is added to *α*_*c*_ this choice corresponds to the date when eq. 1 visually becomes different of the mono-exponential (see supplementary file). *γ* was assumed country independent as we search for an impact of the latitude of its own. This further allows to prevent an over-fitting of the data noise by a steepness tuned for each country.

Note that as the exponential coefficient varies with time, the doubling time around the surge date is not simply ln(2) divided by this coefficient.

### New daily cases dynamics models

In order to evaluate the impact of the UV insolation on the new daily cases dynamics we will consider the simple model:

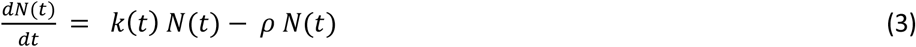

where *N*(*t*) is the total number of persons who have the active sars-cov-2 at time *t*, its derivative being the new daily cases; *k*(*t*) is the mean effective contagiousness of an infected subject which mainly depends on his coronavirus release in air and on materials, on the closeness and frequency of his contacts with other subjects.; *ρ* is the recovery rate. If the contagiousness is constant, the daily new cases follows a mono-exponential increase or decrease:

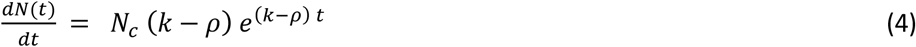

Where Nc is the initial number of infected persons in country c. We will consider two impacts of UV insolation: outdoor sars-cov-2 inactivation and blood 25-hydroxyvitamin D (25(OH)D) concentration.

### Impact of outdoor sars-cov-2 inactivation by solar UV

The active fraction survival of sars-cov-2 *f* under a constant UV insolation R is governed by:

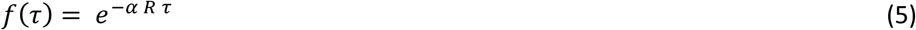

where α is the solar UV sensitivity of sars-cov-2 and *τ* the insolation duration. A recent detailed analysis [2] shows that in Europe the *τ*_*90*_, i.e. the noon solar insolation duration needed to inactivate 90% of the sars-cov-2, approximatively linearly increases between August and October from 60 to 150 minutes for south countries, and from 100 to 250 minutes for north countries.

Considering this inactivation as the single effect varying *k(t)*, we get:

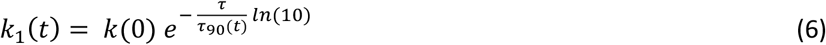

where *τ* is the mean time between the infection of a material and the contact of this material by an non-infected person.

Fig. 2A shows a typical *k*_*1*_*(t)* curve for *τ*_*90*_ ranging from 60 to 250 min, and *τ* = 30 min (*τ* can be modified in the supplementary excel file).

**Figure 2:**
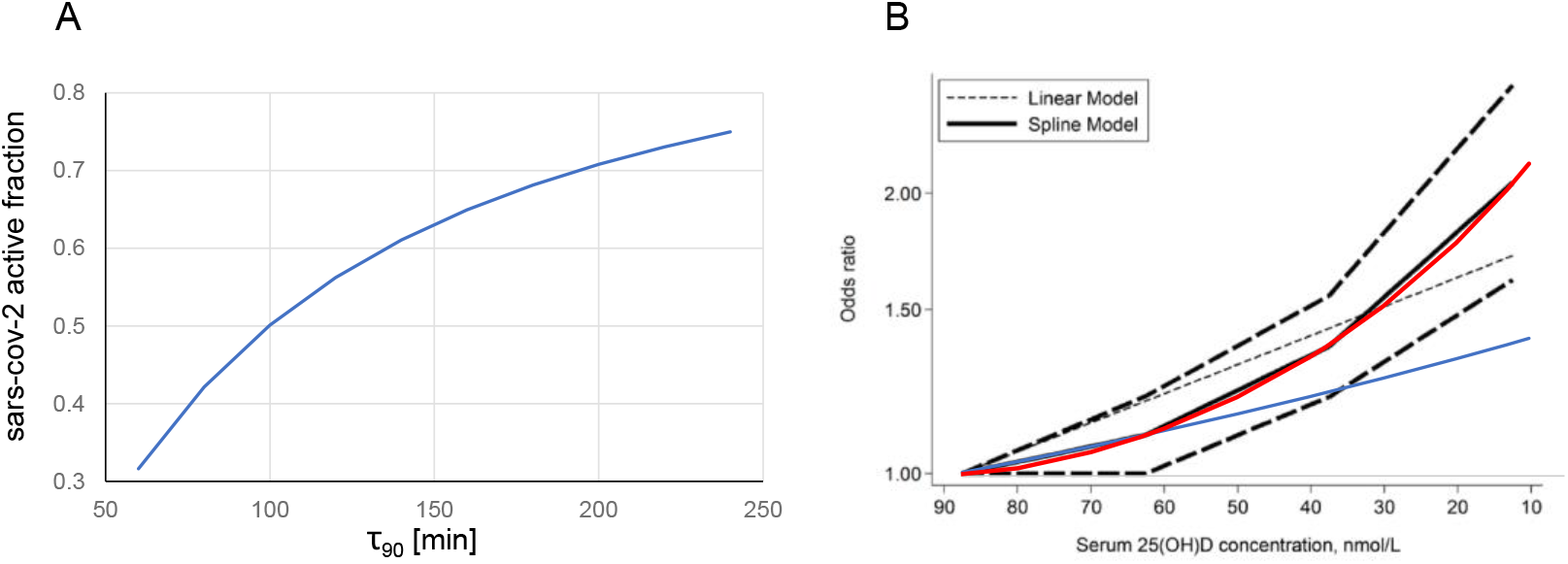
A: survival active sars-cov-2 fraction after 30 minutes solar UV insolation as a function of τ_90_. B: solid black line: acute respiratory track infection (ARTI) risk during cold and influenza epidemy as a function of the 25(OH)D concentration (reprinted from [3] with authorization of ..); red and blue curves: power-exponential (ed. 7) and first point mono-exponential fits added by the present author. In both graphs, corresponding date is running from left to right. Note that the ARTI risk after a while strongly diverges from the mono-exponential.

### Impact of blood (25(OH)D) concentration

A meta-analysis of 24 studies reporting the association between 25(OH)D concentration and risk or severity of acute respiratory track infection (ARTI) during cold or influenza epidemic [3] shows that the risk follows an power-exponential relation (Fig. 2B).

Assuming that covid-19 risk similarly depends on 25(OH)D concentration and considering this risk increase as the single effect varying *k(t)*, we get

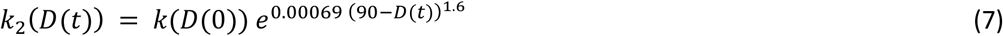

where *D(t)* is the 25(OH)D concentration expressed in nmol/L units at time *t*.

Studies in Europe[4-8] reported a seasonal 25(OH)D concentration drop of 20-26 % between August and October (table 1). Two longitudinal studies [6,7] followed one single cohort during 12 months, with one [7] reporting the 25(OH)D concentration curves for each subjects allowing a rough estimation of the intra-individual standard deviation, giving a drop of 26 ± 25% in normal subjects (age 31 ± 3 years).This standard deviation could still be higher in country population regarding the presence of older, and also of chronically ill, subjects. As a results, more than 15% of European population could from August to October suffer from a 25(OH)D concentration decrease larger than 50%. Assuming that the COVID-19 risk follows fig. 2B and that the whole population has an initial D = 90 nmol/L value, a rough estimation of the k(t) surge can be computed as(see excel sheet “vitD severity” S-AA for the numerical integration result):

**Table 1:** monthly seasonal 25(OH)D concentration studies

| country [ref] | PWC lat. [deg] | population | type | n ± std per month | age [year] | drop ± std [%] |
| --- | --- | --- | --- | --- | --- | --- |
| Sweden [4] | 59.0 | blood donors | ≠ cohorts | $45 \pm 9$ | $41 \pm 13$ | 20 |
| Denmark [5] | 55.9 | blood donors | ≠ cohorts | $16 \pm ?$ | 24-89 | 23 |
| UK [6] | 52.8 | 1958 birth | longitudinal | 6789 | $45 \pm 0$ | $26 \pm ?$ |
| Netherland [7] | 52.1 | nurses | longitudinal | 8 | $31 \pm 3$ | $26 \pm 25$ |
| Poland [8] | 51.7 | athletes | ≠ cohorts | $229 \pm 55$ | $25 \pm 1$ | 23 |

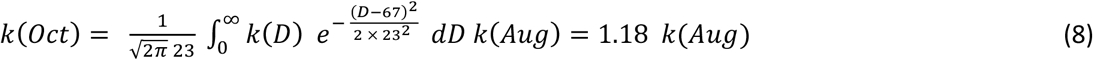

In the integral, 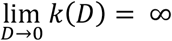 but due to higher exponent power of the gaussian, i.e. 2, versus that of *k(D)*, i.e. 1.6, the integral does not diverge and can even be truncated at *D = 0*. The integration was extended above the initial D = 90 nmol/L value, regarding that one patient in the cohort [7] exhibited a concentration increase rather than a drop.

Right column: 25(OH)D concentration drop between August and November

## Results

Table 2 shows the fitting results (all data and fitting process are provided in the supplementary xlsx file). For Sweden the new daily cases were constant before August preventing the computation of the β/α ratio.

**Table 2.**
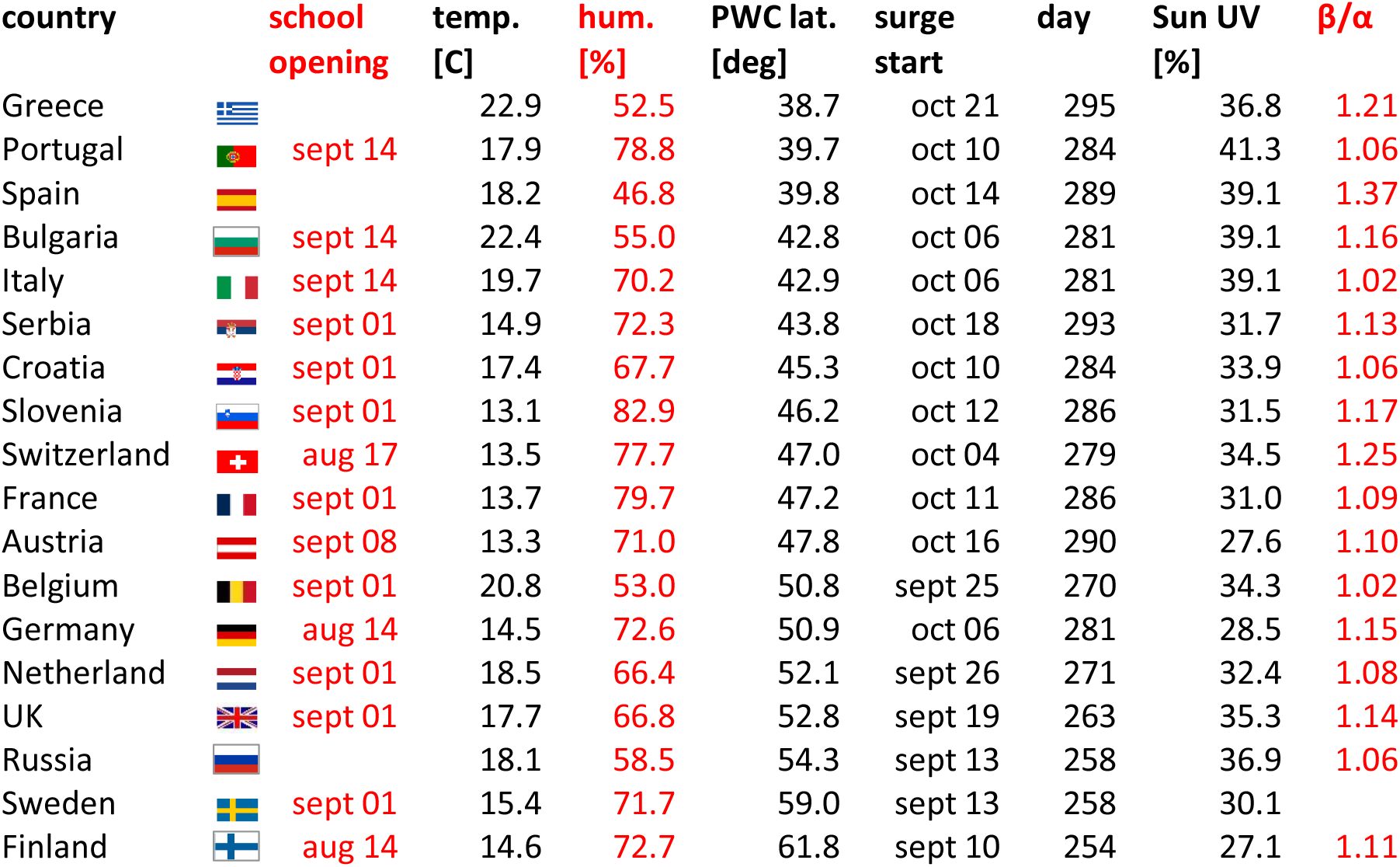
Temp, hum: mean country temperature and humidity during the 2 weeks preceding its COVID-19 surge date. PWC lat: latitude of the country PWC. Sun UV: theoretical sun UV daily dose at the surge date, expressed as a fraction at the PWC latitude versus the latitude 0°.

| country | school opening | temp. [C] | hum. [%] | PWC lat. [deg] | surge start | day | Sun UV [%] | $\beta/\alpha$ |
| --- | --- | --- | --- | --- | --- | --- | --- | --- |
| Greece |  | 22.9 | 52.5 | 38.7 | oct 21 | 295 | 36.8 | 1.21 |
| Portugal | sept 14 | 17.9 | 78.8 | 39.7 | oct 10 | 284 | 41.3 | 1.06 |
| Spain |  | 18.2 | 46.8 | 39.8 | oct 14 | 289 | 39.1 | 1.37 |
| Bulgaria | sept 14 | 22.4 | 55.0 | 42.8 | oct 06 | 281 | 39.1 | 1.16 |
| Italy | sept 14 | 19.7 | 70.2 | 42.9 | oct 06 | 281 | 39.1 | 1.02 |
| Serbia | sept 01 | 14.9 | 72.3 | 43.8 | oct 18 | 293 | 31.7 | 1.13 |
| Croatia | sept 01 | 17.4 | 67.7 | 45.3 | oct 10 | 284 | 33.9 | 1.06 |
| Slovenia | sept 01 | 13.1 | 82.9 | 46.2 | oct 12 | 286 | 31.5 | 1.17 |
| Switzerland | aug 17 | 13.5 | 77.7 | 47.0 | oct 04 | 279 | 34.5 | 1.25 |
| France | sept 01 | 13.7 | 79.7 | 47.2 | oct 11 | 286 | 31.0 | 1.09 |
| Austria | sept 08 | 13.3 | 71.0 | 47.8 | oct 16 | 290 | 27.6 | 1.10 |
| Belgium | sept 01 | 20.8 | 53.0 | 50.8 | sept 25 | 270 | 34.3 | 1.02 |
| Germany | aug 14 | 14.5 | 72.6 | 50.9 | oct 06 | 281 | 28.5 | 1.15 |
| Netherland | sept 01 | 18.5 | 66.4 | 52.1 | sept 26 | 271 | 32.4 | 1.08 |
| UK | sept 01 | 17.7 | 66.8 | 52.8 | sept 19 | 263 | 35.3 | 1.14 |
| Russia |  | 18.1 | 58.5 | 54.3 | sept 13 | 258 | 36.9 | 1.06 |
| Sweden | sept 01 | 15.4 | 71.7 | 59.0 | sept 13 | 258 | 30.1 |  |
| Finland | aug 14 | 14.6 | 72.7 | 61.8 | sept 10 | 254 | 27.1 | 1.11 |

Fig. 3A,B clearly prove no correlation with temperatures or humidity, while fig. 3C clearly shows an impact of the country latitude.

**Figure 3:**
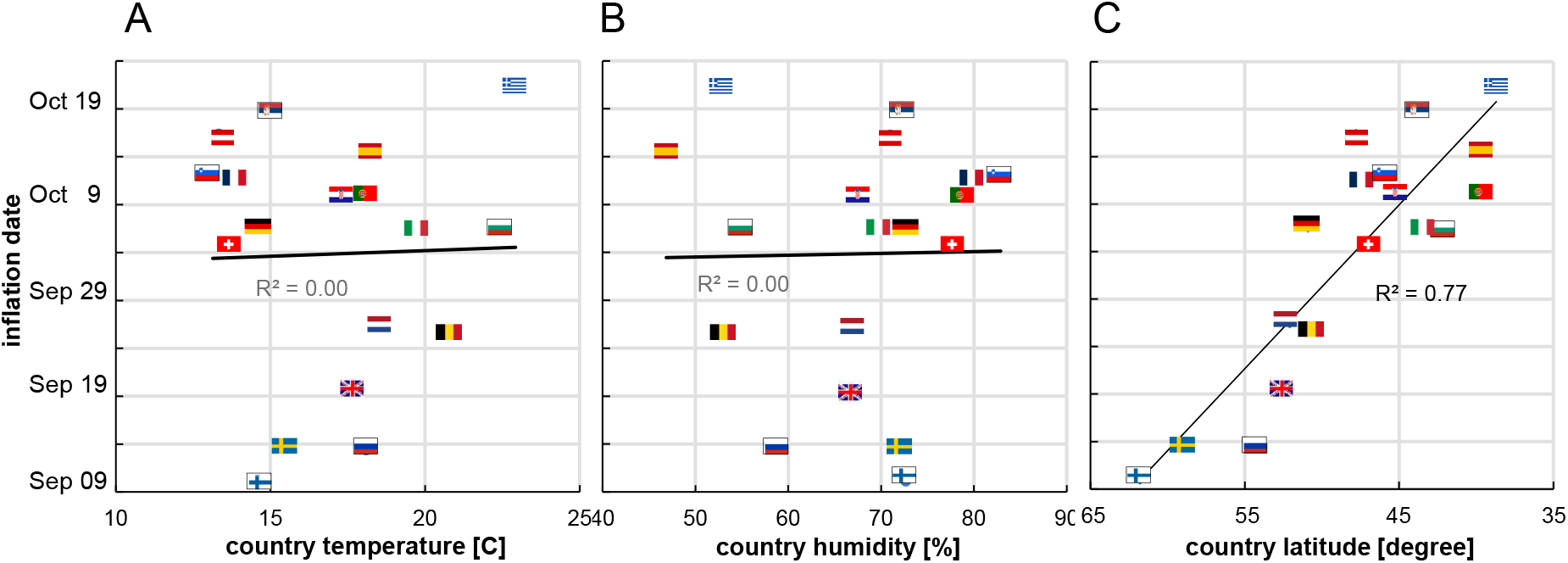
COVID-19 surge date as a function of the country mean temperature (A) and humidity (B) during the 2 preceding weeks, and as a function of the country PWC latitude (C), pointing vitamin D as one of the primary factor (flags link country between graphs).

Fig. 4 clearly shows that the surge dates set on the sun UVB daily dose as a function of the latitude evidences an impact of the sun UVB daily dose.

**Figure 4:**
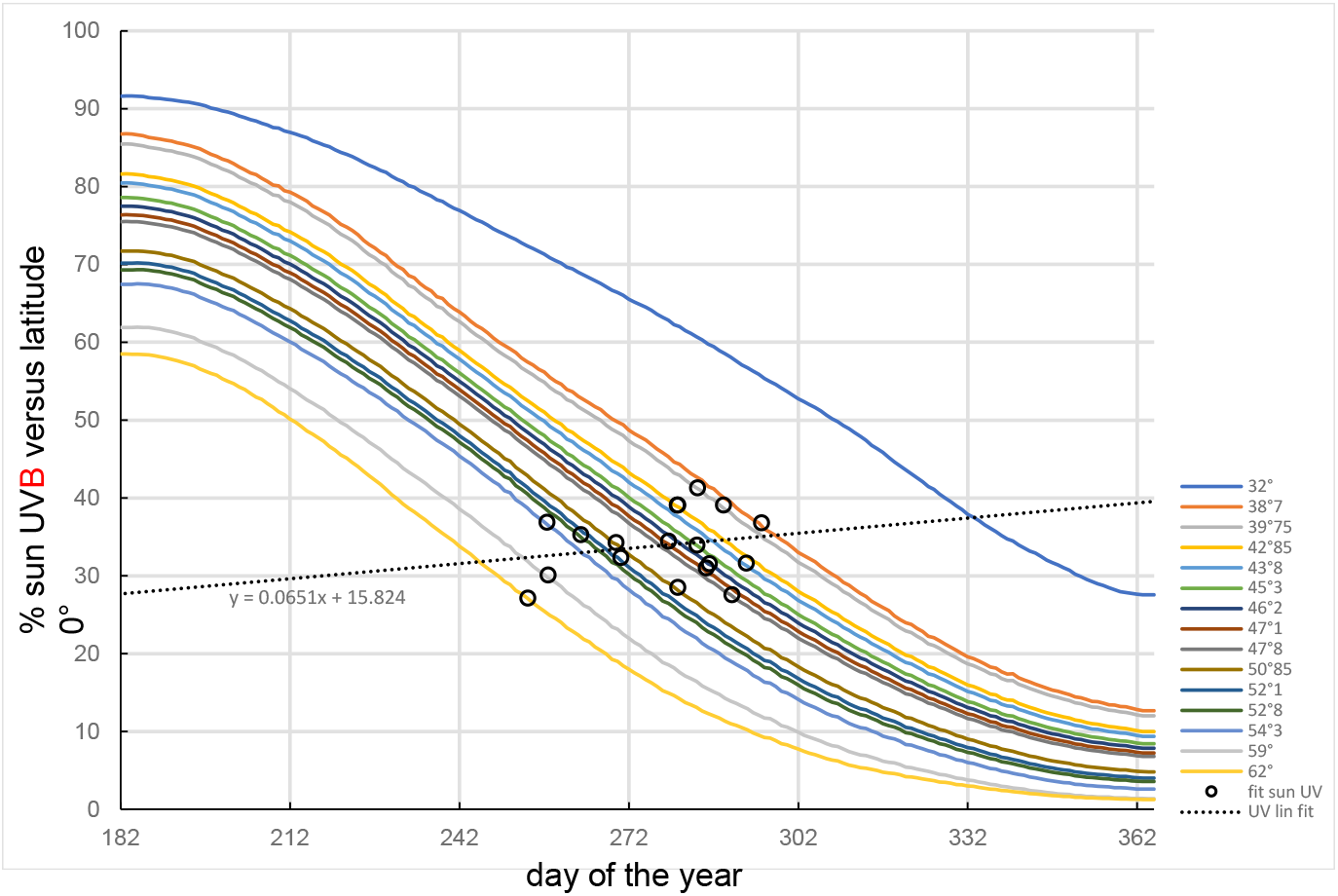
Solid curves: theoretical sun UVB daily dose for vitamin D skin production under a clear sky corresponding to the 18 PWC country latitudes plus the 31° latitude (derived from fig. 1B in [1]). Black circles: country surge dates positioned on their corresponding latitude curve.

Fig. 5 shows that the day of the second wave surge is well predicted by the time when the sun UVB daily dose of the country becomes lower than 30% of that at latitude 0°.

**Figure 5:**
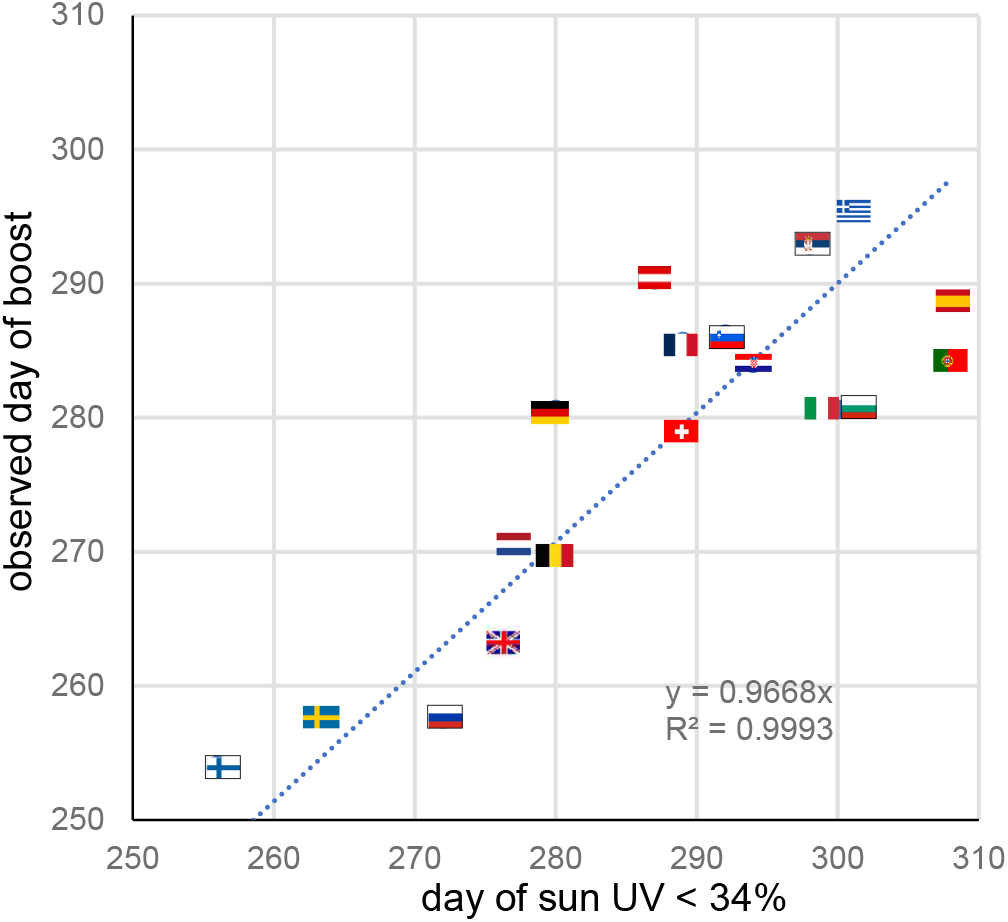
observed day of the second COVID-19 wave surge as a function of the day when the country sun UVB daily dose drops lower than 34% of that at latitude 0°. Trendline forced to intercept (0,0).

## Discussion

Many studies support an impact of low 25(OH)D concentration on the respiratory impairment in coronavirus or virus diseases [9], and also recently for the COVID-19 pandemic (see [10] for a detailed review and analysis of 14 studies reporting such impact). Low 25(OH)D concentration is also more prevalent in populations at risk: i.e. aged people [11,12], obese patients [13], coloured skin people living in high latitude countries [14] and diabetic patients [15].

However by comparing the covid-19 severity between different populations, discriminate whether the 25(OH)D concentration is a real factor of covid-19 severity or only a marker of another weakness being the primary severity factor can be challenging.

The date of the surge is an intrapopulation observation and has the benefit to be only triggered by a parameter globally affecting this population. There is no correlation with temperature, nor with humidity, nor with school opening dates (see excel file), but an impressive latitude correlation (fig. 3). The remaining common parameter affecting these populations at different times monotonously depending on the latitude is the sun UV daily dose (fig. 4).

This UV index dependence was already observed for influenza epidemy [16], but also associated with a temperature dependence. A global seasonality study also evidenced a monthly corelation for other pre-existing human seasonal coronaviruses activity with temperature and humidity [17]. However, this study did not consider the latitude as a confounding factor, and on a monthly scale there is a correlation between temperature-humidity and latitude. On the daily scale used in the present study, this correlation does not longer exist, each country being temporally affected by different wind directions. This feature allows to clearly discriminate between temperature-humidity and latitude impact.

The decreasing sun UV insolation can impact the covid-19 dynamics in two ways: by decreasing the outdoor sars-cov-2 inactivation or by decreasing the population 25(OH)D concentration.

However, many European countries were able in November to break the surge by additional safety rules. Activities where people cannot wear the face mask such as collective sport, or such as relaxing in pubs and restaurants were forbidden, also festive activities where people often forget distancing recommendations were forbidden. In contrast, population continues their professional and outdoor relaxing activities wearing the face mask at work, in public transport, in itinerant outdoor markets (an European use) and in parks. The success of these rules supports the major role of airborne transport of the sars-cov-2 versus contamination by outdoor infected material contact.

Eq. 6,7 clearly illustrate that the potential impacts on covid-19 dynamics of outdoor sars-cov-2 inactivation and of 25(OH)D concentration decreasing are fundamentally different. Indeed, Fig. 2B supports that even if the 25(OH)D concentration slowly decreased after the summer solstice, its impact on the contagiousness becomes more and more important with time and leads the dynamics to strongly diverge from a mono-exponential after a while, this is in line with the occurring in October. In contrast, Fig. 2A supports that the impact of the outdoor sars-cov-2 inactivation decreasing on the covid-19 contagiousness becomes less and less important with time, which should had correspond to a enhance in July-September, moving towards a doubling time stabilization.

The obtained β/α ratios range from 1.02 to 1.37 (Table 2), this is in line with the estimated ratio 1.18 regarding that eq. 8 neglects any country dependence and was based on a small cohort (n=8) follow up.

The positive linear slope of the sun UVB threshold versus the country latitude (Fig. 4) is also in line with the fact that, due to the natural adaptation, populations have a more and more pigmented skin when the latitude decrease. As a result, skin vitamin D production in north populations is affected by the sun UVB decrease in a slower way than that of the south populations. Fig. 4 is also in line with the low population mortality observed within ±35° latitudes [18] and reported in Hubei located on the 31° latitude, regarding that these regions are most of the year above the sun UV daily dose 34% average threshold.

The present study supports thus that low 25(OH)D concentration is well a contributing factor of COVID-19 severity as already showed by the previous studies [10], which all together constitutes a convincing bundle of evidence. By increasing the coronavirus load in the respiratory track, the contagiousness in the population is also increased, starting a chain reaction which explains the wave surge.

## Conclusion

As already evidenced by previous correlation studies [10], low 25(OH)D concentration should be considered as a contributing factor of covid-19 severity.

Europe, and north USA as well, are starting this autumn a long covid-19 crisis as they will get back above the October sun UV daily dose only end 2021 March.

The bet to reduce the pandemic severity during the coming winter using controlled preventive vitamin D supplementation should be considered [10, 19].

## Supporting information

data and fit excel file

## Data Availability

all data referred in the paper come from open database

