## Supplementary material for "The autumn COVID-19 surge dates in Europe are linked to latitudes and not to temperature, nor to humidity, pointing vitamin D as a contributing factor": excel file info

corresponding author:

**Excel file: data fitting**

In each country page it is possible to visually localize the surge date, i.e. by changing the day value in the blue cell, the corresponding correlation graph is also added in the RESULTS sheet just below the automatic one. The temperature-humidity averaging are in columns S-AA. The mono-exponential curve (thin black curve) is also displayed on the daily new cases data.

The manual correlation is similar than the automatic one, i.e. $R^2$ = 0.75, but for some countries with noisy data, it is not easy to visually determine the surge date. This noise mainly results from the poor reporting performed during the WE, i.e. the Saturday and the Sunday.

For Spain and Switzerland (d>262), the Saturday, Sunday and Monday (column Q) cases were reported together to the Monday, so I split the Monday cases in three to have an useful curve. But for many countries it is like a random part of the WE daily cases were lost.

There is a mean offset of 7.2 days (pink cell in RESULTS sheet) between automatic and manual determination, it is like eye see the surge when 11 % of (β-α) is added to α, and not 50% as arbitrary stated in the paper. However, it is possible to also use 11% for the automatic fit by putting 7.2 in the blue cell in the RESULTS sheet (current excel state), obviously this has no impact on the correlation goodness.
